# DEPOT: graph learning delineates the roles of cancers in the progression trajectories of chronic kidney disease using electronic medical records

**DOI:** 10.1101/2023.08.13.23293968

**Authors:** Qianqian Song, Xiang Liu, Zuotian Li, Pengyue Zhang, Michael Eadon, Jing Su

## Abstract

Chronic kidney disease (CKD) is a common, complex, and heterogeneous disease impacting aging populations. Determining the landscape of disease progression trajectories from midlife to senior age in a real-world context allows us to better understand the progression of CKD, the heterogeneity of progression patterns among the risk population, and the interactions with other clinical conditions like cancers. In this study, we use electronic health records (EHRs) to outline the CKD progression trajectory roadmap for the Wake Forest Baptist Medical Center (WFBMC) patient population. We establish an EHR cohort (n = 79,434) with patients’ health status identified by 18 Essential Clinical Indices across 508,732 clinical encounters. We develop the DisEase PrOgression Trajectory (DEPOT) approach to model CKD progression trajectories and individualize clinical decision support. The DEPOT is an evidence-driven, graph-based clinical informatics approach that addresses the unique challenges in longitudinal EHR data by systematically using the graph artificial intelligence (graph-AI) model for representation learning and reverse graph embedding for trajectory reconstruction. Moreover, DEPOT includes a prediction model to assign new patients along the progression trajectory. We successfully establish the EHR-based CKD progression trajectories with DEPOT in the WFUBMC cohort. We annotate the trajectories with clinical features, including kidney function, age, and other indices, including cancer. This CKD progression trajectory roadmap reveals diverse kidney failure pathways associated with different clinical conditions. Specifically, we have identified one high-risk trajectory and two low-risk trajectories. Switching pathways from low-risk trajectories to the high-risk one is associated with accelerated decline in kidney function. On this roadmap, high-risk patients are enriched in the skin and GU cancers, which differs from low-risk patients, suggesting fundamentally different disease progression mechanisms. Overall, the CKD progression trajectory roadmap reveals novel diverse renal failure pathways in type 2 diabetes mellitus and highlights disease progression patterns associated with cancer phenotypes.

## Introduction

Chronic kidney disease (CKD) is common, affecting 14.8% of US adults and disproportionately more in diverse and underserved communities [1]. CKD significantly reduces life expectancy and quality of life while imposing a tremendous economic burden on society. The progression of CKD is multifactorial with highly heterogeneous disease trajectories, reflecting long-term risk factors from lifestyle, diet, nephrotoxic medication use, genetic traits, socio-economic determinants, and other medical comorbidities. As a chronic, complex, and heterogeneous disease, CKD challenges traditional clinical research paradigms such as those in clinical trials or observational studies. One major analytic challenge is the heterogeneity in the CKD population that demonstrates dramatically diverse progression patterns, prognosis, risk factors, and responses to clinical interventions. The other challenge is the complexity of the CKD population which requires comprehensive clinical indices to study its risk of progression, interactions with its comorbidities like cancers, and the impact on health outcomes. As CKD is often diagnosed by estimated glomerular filtration rate (eGFR) [2-4], a critical need persists for identifying the modifiable risk factors in susceptible populations early and establishing actionable support for medical decision-making.

Electronic health records (EHRs) provide unique and valuable opportunities to address these challenges in clinical decision support for managing the risks of CKD. EHRs cover large cohorts over decades from various aspects, including demographic, clinical, financial, and socioeconomic features. Modern advances in big data science significantly improve data interoperability, making meta-analysis in cross-regional or national EHR networks possible. Thus, an EHR-based clinical decision support system has the potential to revolutionize the care of patients with CKD. However, the challenges of using EHR-related big data have impeded longitudinal studies of CKD progression. Although the EHR provides rich and comprehensive information about the population across a wide age span, for a specific clinical visit, or an individual patient, EHR data are often sparse with irregularly spaced intervals. These factors challenge traditional approaches that require data completeness and regularity. The recent advance in topological learning on big data, especially novel graph learning algorithms [5-8], allow building the trajectory tree according to the similarities among data points from highly scattered data, thus can be used to outline the common disease progression trajectories of a population using the EHR data as a whole. This strategy enables unleashing the power of EHR data and overcoming traditional approaches’ limitations.

In this study, we focus on the high-risk population with type 2 diabetes mellitus as the study cohort, linking participants from a series of genomics studies with their EHRs at Wake Forest Baptist Medical Center (WFBMC), using EHR data in 79,434 patients, including 18 essential clinical indices, 508,732 clinical encounters, and clinical records to learn and annotate the declines in chronic health condition trajectories during aging. The WFUMBC EHR data are high-quality and research-ready after data harmonization, curation, and quality control by the Clinical and Translational Sciences Institute. However, challenges persist in analytical and modeling approaches on such heterogeneous and complex data, including 1) data fragmentation, 2) limited structural data availability, 3) generalizability of learned knowledge due to variability across health systems, and 4) various temporal patterns in clinic visits, treatment outcomes, and follow-up, which reflect the diversity of patient experience. Novel strategies are required to fulfill the promises of using longitudinal EHR data to identify and predict CKD progression patterns precisely.

To address these challenges, we propose to develop the DisEase PrOgression Trajectory (DEPOT), an evidence-driven, graph-based clinical informatics approach to model CKD progression trajectories and individualize clinical decision support. The graph-based DEPOT approach can address the unique challenges in longitudinal EHR data by using the graph-AI model for representation learning of temporal patterns of clinical encounters and the reverse graph embedding for trajectory reconstruction. We hypothesize that there are different CKD progression paths which are: 1) driven by different pathogenic mechanisms, 2) susceptible to different nephrotoxic drugs, and 3) identified by unique EHR data patterns. Mathematically, such CKD trajectory landscapes can be learned as principal graphs representing the topological and temporal characteristics of the observed, fragmented EHR data. With the DEPOT approach, we have established EHR-based CKD progression trajectories, examined the chronic renal function declines on these trajectories, and highlighted those closely associated with cancer patients. We also examine the predictive power of the learned disease progression trajectories. Our identified results cast new light on understanding the diversity of CKD progression paths and their associated health conditions.

## Data and Methods

### The WFBMC EHR cohort

The WFBMC EHRs are composed of the pre-Epic “Legacy” EHRs (1985 – 2012), providing invaluable longitudinal records for chronic disease research, and the Epic-based “WakeOne” EHRs (since 2012), providing modern and fast-growing EHRs for clinical research as well as the major implementation platform for delivering clinical decision support. The WFUHS’s Informatics for Integrating Biology & the Bedside (I2B2)-based Translational Data Warehouse (TDW) integrates both Legacy and WakeOne EHRs, implements a wide range of biomedical ontologies and terminologies for EHR presentation and annotation, and curate a high-quality data warehouse for biomedical and clinical research. The WFUHS Center of the Scalable Collaborative Infrastructure for a Learning Health System (SCILHS), the National Patient-Centered Clinical Research Network, an innovative initiative of the Patient-Centered Outcomes Research Institute (PCORI), has established a comprehensive and cutting-edge EHR system according to Unified Medical Language Systems (UMSL) as prescribed in the PCORNET Common Data Model (CDM) (www.pcornet.org/resource-center/pcornet-common-data-model) to support large scale clinical informatics and phenome mining. The working cohort included 79,434 type 2 diabetes-affected patients and their EHR data between 1993 and 2018.

### CLINIC CDM

We curated the *Common cLINic <u>I</u>ndex for Chronic diseases* (CLINIC) CDM to comprehensively outline the general health conditions during the development and progression of common chronic diseases. The CLINIC CDM is composed of the following categories:

- Demographics (3 features): date of birth (DoB), sex, self-reported race;
- Vitals (5 features): diastolic and systolic blood pressure, height, weight, and body mass index (BMI);
- Laboratory tests (14 features): alanine aminotransferase (a.k.a. serum glutamic pyruvic transaminase), aspartate aminotransferase (a.k.a. serum glutamic oxaloacetic transaminase), alkaline phosphatase (ALK), total cholesterol, low-density lipoprotein cholesterol (LDL), and high-density lipoprotein cholesterol (HDL), creatine kinase, estimated glomerular filtrating rate (eGFR), hemoglobin, hemoglobin A1c (HbA1c), triglycerides, international normalized ratio of prothrombin time (INR), serum creatinine, total bilirubin (TBIL), and troponin;
- Diagnosis: ICD9 and ICD10 codes;
- Procedures: HCPCS, ICD9-CM, and ICD10-PCS codes;
- Medications: RxNorm codes.

The BMI was derived from height and weight. The eGFR was estimated according to serum creatinine, age, sex, and race with the CKD-EPI Creatinine Equation (2009). The CLINIC CDM is compatible with Carolinas Collaboratives Common Data Model (CDM) and PCORI CDM.

### The Essential Clinical Indices (ECI) of clinical encounters

We defined an 18-feature Essential Clinical Indices for each clinical encounter to quantitatively describe the overall health conditions of patients. These indices comprise all vital and laboratory variables in the CLINIC CDM except BMI since BMI was highly correlated with height and weight. The selection of these clinical indices was a tradeoff of the following considerations:

- Reflect on health status and risks of chronic disease with aging, especially those associated with CKD risks.
- Clinically measured, providing strong evidence.
- Generally available in EHR.
- Readiness to use. Most of these indices had already been cleaned and aggregated by CTSI at WFBH as part of the Carolinas Collaborative infrastructure.

### Cancer diagnosis and phenotypes

We use a similar primary cancer diagnosis algorithm as we previously reported[9, 10]. Briefly, patients with cancers are identified by the 9^th^ and the 10^th^ revisions of the International Statistical Classification of Diseases and Related Health Problems (ICD-9 and ICD-10). The apparent cancer onset date, as well as the primary cancer type for each cancer patient, is estimated by the first diagnosis. Totally 11 major cancer types are phenotyped, including female breast cancer (BR), endocrine cancer (Endo), female reproductive system cancers (excluding breast cancers), gastrointestinal cancers (GI), genitourinary cancers (GU), hematological cancers (Heme), cancers of the neurologic system (Neuro), male reproductive system cancers (excluding prostate cancer), prostate cancer (PRAC), skin cancers (Skin), and thoracic cancers (Thor). Other cancers with known origins are grouped as Others. Cancers of unknown, unspecified, or undocumented origins are labeled as NO.

### EHR data aggregation

The laboratory tests in WFBMC EHR were extracted from both WakeOne (WFBMC’s implementation of the Epic system) and pre-Epic systems. Therefore, a clinical feature is often referred to by different names in the EHR. The original laboratory test names were manually harmonized and aggregated to the CLINIC CDM.

### Graph-based representation learning

In DEPORT approach, we adapted a graph-based artificial intelligence (graph-AI) model, GraphSAGE (SAmple and aggreGatE) [11], for representation learning to address challenges of machine learning on longitudinal EHR and achieve reliable trajectory learning. This graph-AI model embedded the heterogeneous, informatively censored, and mutually dependent EMR data with irregularly longitudinal intervals into a latent space of desirable mathematical properties for trajectory learning.

The original longitudinal EMR data was first represented as graph-structured data. We constructed the age graph of clinical visits to capture the intrinsic patterns of clinical features shared by patients at similar ages. In an age graph, each node represents a clinical visit, and each edge links two clinical visits of similar ages. Specifically, a *k* −*dimensional*tree was used to identify approximate *k*-nearest neighbors (*k*=100) according to the ages of encounters. Each identified clinical visit pair was further screened according to the age difference with a cutoff of 1 month. Demographic and clinical features at each clinical encounter (node) were represented as the attributes of the corresponding node. Data availability patterns in each clinical or demographic feature provide crucial health information. Such informative censoring patterns were captured by specific variables. For example, a feature “eGFR availability” was created to capture whether eGFR values are available in encounters. Meanwhile, the missing values in the original eGFR variable were filled with median eGFR values, and correspondingly, a rectified linear unit (ReLU) was used as the activation function in the GraphSAGE model. In this way, the filled values have minimum impact on the graph AI representation. A patient may have multiple clinical visit records. The pattern of irregular intervals among these visits carried information on medical needs, access to healthcare, and personal lifestyle. The interval between two adjacent clinical visits was also used as an attribute of the corresponding nodes. In summary, the age graph captured the essential topological patterns in longitudinal EMR data and supports learning embedding models according to clinical observations of similar age.

The graph embedding was learned using GraphSAGE with a mean aggregator. Briefly, the input of the graph AI model was the age graph *G*(*V; E*) and the input features {***x***_*v*_, ∀*v* ∈ *V*}, where *v* represents a node (clinical visit). The information of the original graph-structured data was propagated to the latent space through *K* neuron network layers, with the *k* ‘th propagation function (*k* ∈ {1, ⋯, *K*}) as:

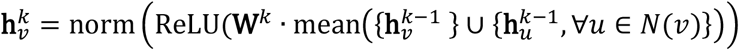

where *u* represents one of *v*’s neighbor noda a a es *N*(*v*) (that is, another clinical visit at similar age of visit *v*), 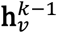 and 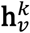 are the associated attributes of node *v* before and after the *k* ‘th information propagation, respectively, **W**^*k*^ is a shared and trainable weight matrix, 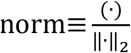 denotes the normalization function, and ReLU indicates the rectified linear unit activation function. The unsupervised, inductive representation of the original features 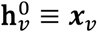 of clinical visit *v* in the latent space is denoted as 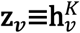.

The graph-based loss function is used for training the unsupervised representation of the heterogeneous raw data in the latent space:

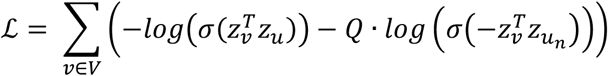

where *u* is a node that co-occurs near *v* on fixed-length random walk, *σ* is the sigmoid function, 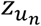 is a non-neighbor node sampled from the negative sample distribution, and *Q* defines the number of negative samples. In the lost function, the first component encourages that clinical visits at similar age are more likely to have shorter distance in the latent space, while the second component keep visits at different ages far away in the latent space.

The related hyperparameters were fine-tuned according to the robustness of the latent representation and the learned trajectories. We performed a parameter sweep on initial learning rates, including 0.01, 0.001, and 0.0001, and selected the optimal one. For the number of hidden units, we checked 9 and 18 in each layer and selected the optimal one. The graph-AI model was implemented in TensorFlow[12] with the Adam[13] optimizer. For all the GraphSAGE variants, we used rectified linear units as the non-linearity.

A random sampling approach (without replacement) was then used to train graph AI models, learn latent representation on very large graphs, and examine the robustness of the results. The study cohort is randomly split into 7 sub-cohorts, each containing 79,434 patients and 72,676 corresponding encounters (nodes). For each sub-cohort (training cohort), a GraphSAGE embedding model is trained, and the embedding of the rest 6 sub-cohorts (predicting cohort) is then predicted using the trained GraphSAGE model. Totally 7 sets of embedding results are calculated for each clinical encounter. The longitudinal and CKD progression patterns were well represented in the latent space.

### Learning disease progression trajectories

The disease progression trajectories were revealed as an accompanied principal graph of the AI-derived latent representation of clinical visits using a discriminative dimensionality reduction tree (DDRTree)[7]. Let 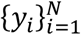 be a set of the *D*-dimensional column vectors in the representation space corresponding to encounters of patients, where *N* is the total number of encounters. We define the latent points as 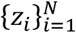 such that *z*_*i*_ is the *d*-dimensional lastent counterpart of *y*_*i*_. To transform *z*_*i*_ to *y*_*i*_, a linear projection matrix *W* ∈ ℝ^*D*×*d*^ is used with *d* ≤ *D*. Moreover, in order to model the discriminative information via clustering, a set of latent points 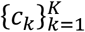 are introduced as the centroids of the latent points 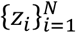 where *c*_*k*_ ∈ ℝ^*d*^. The tree structure is represented by a connectivity indicator matrix *S* between cluster centroids with the (*k, k*′)th element as *S*_*k, k*′_ that is, *S*_*k, k*′_ = 1 means that the *k*-th centroid and the *k*′th centroid are connected in the graph, and 0 otherwise. Specifically, a set of tree structures are denoted by 𝒮_*T*_. By combining the above ingredients, DDRTree is formulated as the following optimization problem

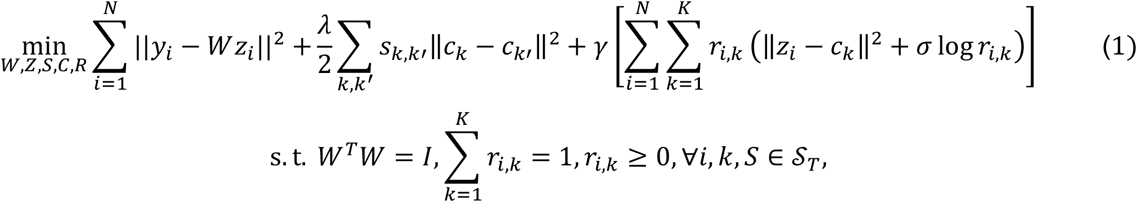

where the first term projects data into a low-dimensional space, the second term is responsible for structure learning, and the third term is interpreted as the objective function of soft K-means [14]. Three hyper-parameters are added to connect the three terms: *λ* ≥ 0 is used for the reversed graph embedding, *γ* ≥ 0 is used o balance the contribution of deterministic clustering, and *σ* ≥ 0 is to regulate the negative entropy regularization. Matrix *R* ∈ ℝ^*N*×*K*^with the (*i,k*)th entry as *r*_*i, k*_ can be interpreted as the probability of assigning *z*_*i*_ to cluster *c*_*k*_.

## Results

### Graph-based models for the deep representation and trajectory learning of heterogeneous EHR data

As demonstrated in Supplementary Figure 1, we first harmonize the longitudinal EMR data collected from Atrium Health Wake Forest Baptist and establish the CKD Phenome data warehouse according to the CLINIC CDM. The Essential Clinical Indices of clinical encounters of the Atrium Health Wake Forest Baptist are used to identify a CKD study cohort composed of 79,434 patients. Then, the longitudinal clinical information, including the informative data availability patterns, is captured as an age graph-structured format. Briefly, an age graph is constructed, with each node representing an encounter (usually a clinical or lab visit), each edge linking two encounters (either from the same patient or from different patients) of similar ages, and essential clinic indices as well as the longitudinal visiting patterns and the data availability patterns of each patient are used as node attributes. After that, we use a graph AI model to capture and represent the rich longitudinal information in the original EHR in a CKD-relevant low-dimensional latent space. Specifically, a customized graphSAGE model [11] is used to learn a graph embedding of the original EHR data into a latent space in which encounters associated with similar original EHR features are located close to each other. Based on the learned graph AI latent representation, we further reconstruct the CKD progression trajectories using a fast and robust reversed graph embedding approach, DDRTree [7]. Finally, the roles of cancers in the discovered CKD trajectories are investigated using association analysis. Two discovered trajectories that impact similar age groups but show different progression patterns are further analyzed.

### Demographical characteristics

The demographic characteristics of the study cohort are summarized in Table 1. The EHR data in the 79,434 patients has rich records and features, including eGFR and cancer information covering 508,732 encounters. There are a total of 922,698 eGFR data, which is the most abundant feature that covers longitudinal visits. In our trajectory learning cohort, we randomly picked 10,000 encounters of 8,402 patients and the other 498,732 of 71,032 patients as the prediction cohort. Across the overall study cohort, the median age of patients is 60.1yr. Based on the categories of solid and liquid cancer, we find that patients with solid cancer (67.7yr) live longer than those with liquid cancer (64.7yr). Females (50.04%) and males (49.96%) are almost equally represented in this cohort. Interestingly, females are shown less represented in both solid and liquid cancer categories. Regarding race and ethnicity, white people mostly dominate (72.20%) the whole cohort, compared with the African American people (22.97%). The ratio of white people versus African American people is comparable with the population distributions in North Carolina. Not surprisingly, this ratio remains similar when looking into the categories of solid cancer or liquid cancer.

### Graph representation and progression trajectory

The graph-AI model learned representation of the EHR-derived ECI of 508,732 encounters from all patient cohorts is shown in Supplementary Figure 2A. Figure 2A shows the representation space with annotations of age, eGFR, race, and sex information. Encounters with elder ages are demonstrated with clear and separate patterns with young ages. For those with older ages, eGFR is mostly lower than those with younger ages, indicating that elder patients have fewer healthy kidney functions than younger patients. Meanwhile, race and sex are organized, suggesting that the representation space is well-learned. Figure 2B presents the learned trajectory based on the representation space. These trajectories are shown in a latent graph space as a tree denoting the gradual development of clinical conditions. Encounters on the trajectory tree are clustered into 30 classes using k-means according to their locations on this tree (Supplementary Figure 2B). These encounters are then annotated by patient age, eGFR, race, and sex information (Figure 2B). Specifically, the eGFR-annotated trajectory tree indicates that these trajectories capture the gradual deterioration of kidney function and disease progression. Moreover, the progression of kidney disease along the trajectories is consistent with the aging directions. Notably, the trajectory tree has multiple progression directions, reflecting the diversity of kidney disease.

**Figure 1.**
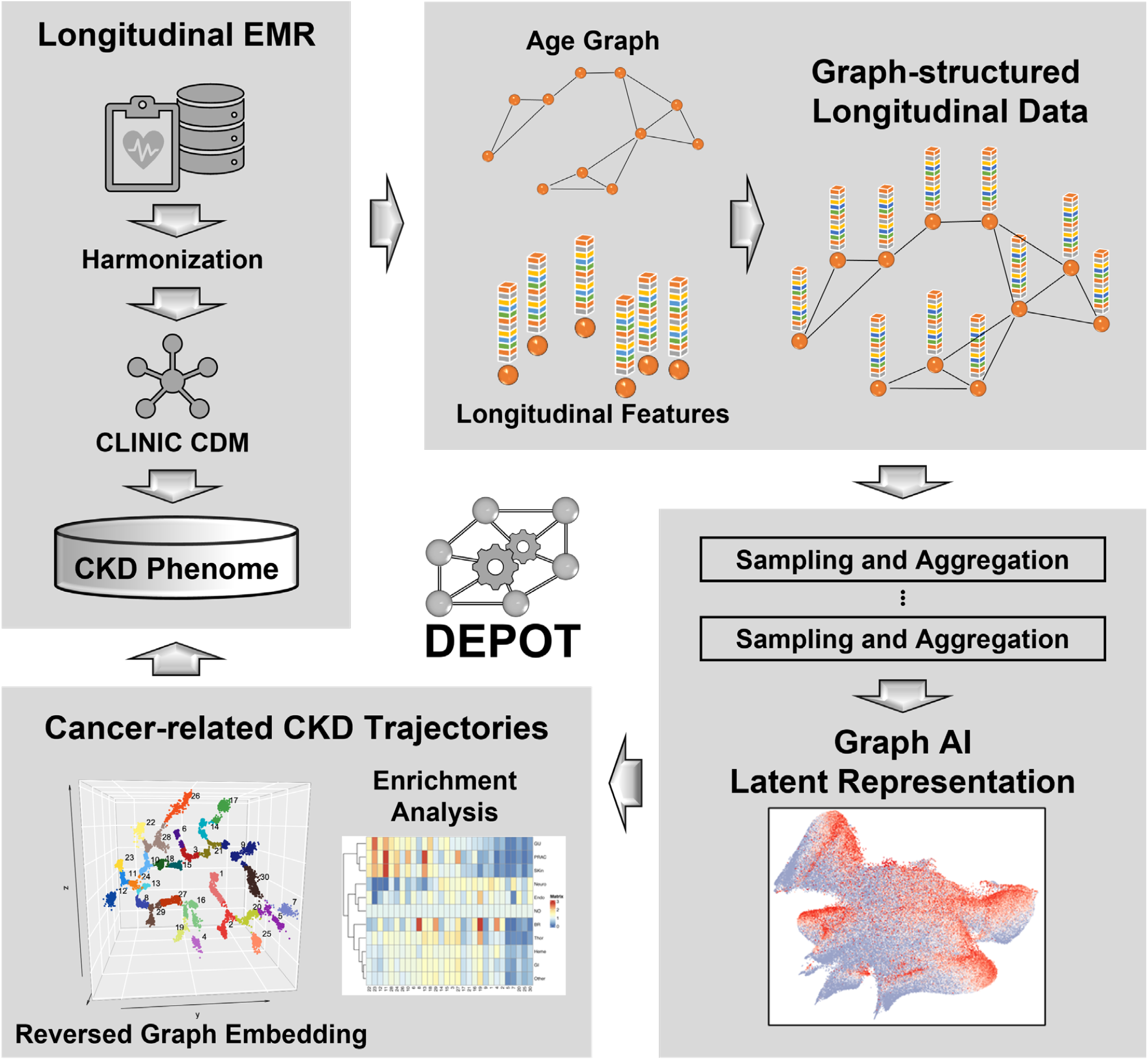
Schematic overflow of DEPORT approach. DEPORT is a graph-based approach for deep representation and trajectory learning of heterogeneous EHR data. In DEPORT, the graph-AI model is first used to capture and represent the rich longitudinal information in EHR in a CKD-relevant low-dimensional latent space. With the learned representation, reversed graph embedding model is then applied to learn disease progression trajectories and predict new encounters.

**Figure 2.**
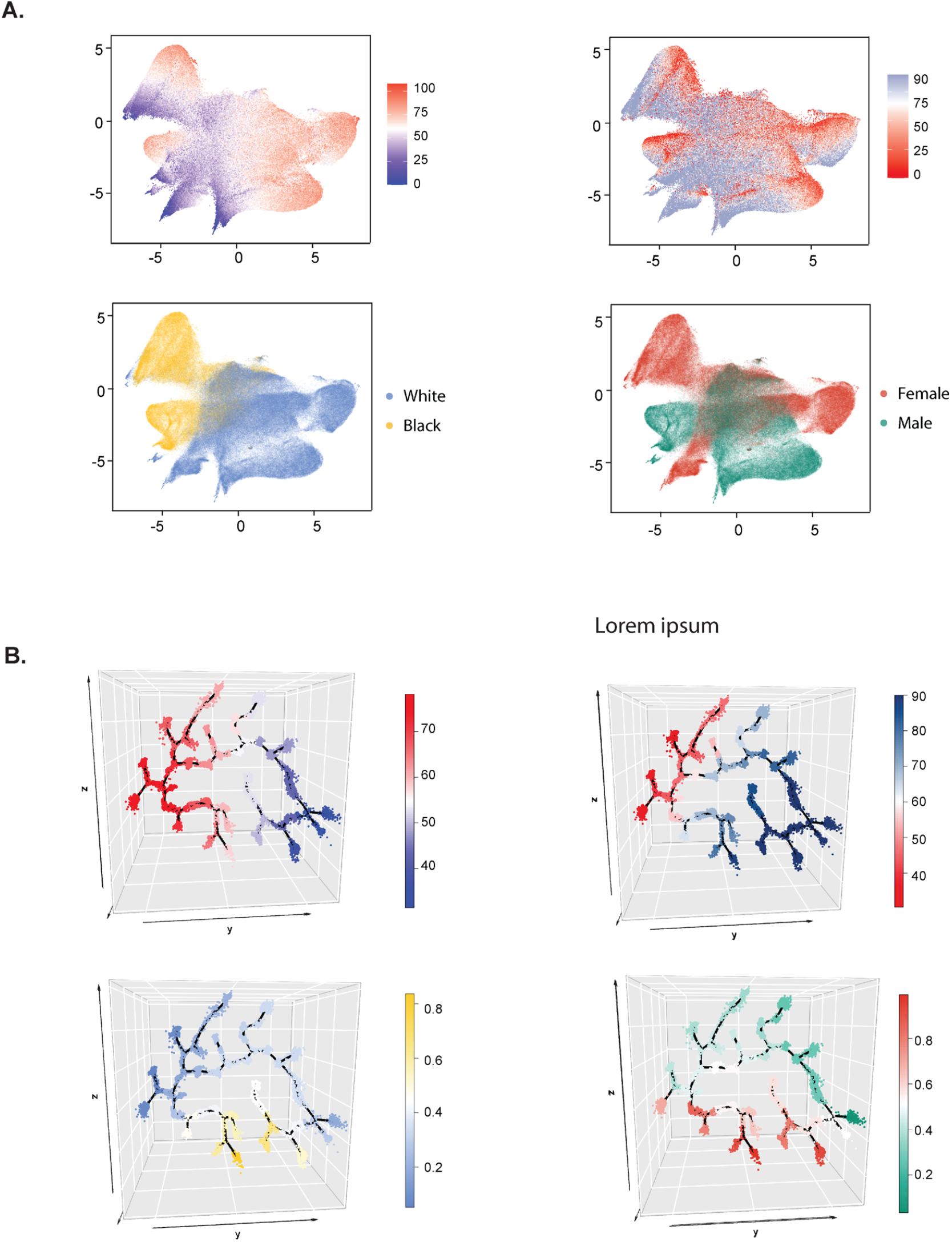
DEPORT identified representation and trajectories. (A) UMAP visualization of the representation space learned by the graph-AI model in our DEPORT approach. The representation space consists of encounters shown as dots and colored coded by their respective age, eGFR value, race, and sex, respectively. (B) Progression trajectories learned from the representation space. Encounters are shown as dots, color-coded by median age, eGFR value, race, and sex, respectively, of corresponding classes.

With the clustered 30 classes of encounters on the trajectory tree (Supplementary Figure 2B), as well as their annotated age, eGFR, race, and sex information (Figure 2B), we then order these classes on the trajectory tree by their ages (Supplementary Figure 2C). Class 12 and 11 are shown as the eldest, while classes 7 and 5 are the youngest. We further investigate the eGFR values across the ordered age classes (Supplementary Figure 2D). Most elder classes (e.g., 12, 11, 23, 22) have lower eGFR values, indicating severe kidney impairment in aging patients. Interestingly, there are also elder classes that have higher eGFR values (e.g., 22, 28), and some young classes have relatively lower eGFR values (e.g., 8, 13), indicating that some older patients also have healthy kidney functions. At the same time, young patients may also have kidney impairment. Specifically, we observe two trajectories; one is the “high-risk” trajectory composed of classes 4, 19, 16, and 27, mostly young people but with lower eGFR values and severe kidney impairment. The other one is the “low-risk” trajectory composed of classes 22, 28, and 26, with older people with higher eGFR values and healthy kidney functions. This suggests that the severe renal functional change in the high-risk trajectory is independent of age. Besides, the proportions of different sex (Supplementary Figure 2E) and races (Supplementary Figure 2F) are evenly distributed in most aging classes.

### Chronic trajectories in patients with cancers

Besides the annotations above, we further identify novel cancer-related CKD progression patterns using term frequency-inverse document frequency [15] (TF-IDF)-based enrichment analysis across all classes on the trajectory tree. With the enriched results (Figure 3A), Neuro cancer appears to dominate more courses than other cancer types. Skin cancer tends to increase in classes with low eGFR values, indicating these skin cancer patients mostly have kidney impairment. Classes with high eGFR values and healthy kidney functions have no enrichment of cancers. To quantitatively delineate different cancer enrichment versus kidney function, we present each cancer enrichment in the heatmap, with the trajectory classes ordered by their respective eGFR values (Figure 3B). Here we have removed the potential confounding effects of age. The heatmap, without eliminating the confounding effects of age, is shown in Supplementary Figure 3. GU, PRAC, and skin cancer are more enriched in lower eGFR classes on the trajectory tree. In contrast, Neuro, Endo, and no cancers are more enriched in higher eGFR classes on the trajectory tree. Interestingly, BR cancer is enriched in two divergent classes with both low and high eGFR values.

**Figure 3.**
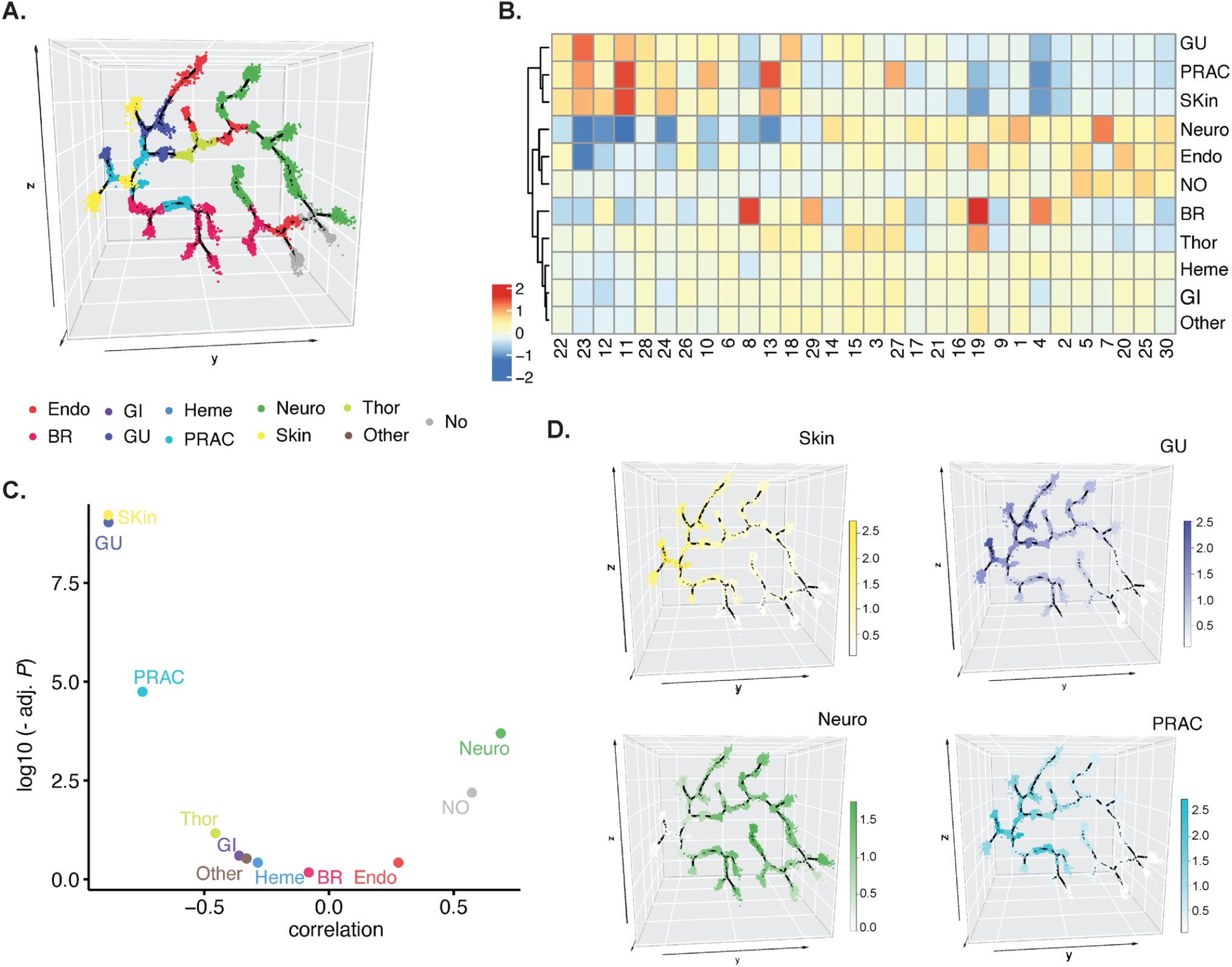
Enrichment of different cancer types on disease progression trajectories. (A) Progression trajectories with encounters color-coded by enriched cancer types. TF-IDF is used to evaluate cancer enrichment in each class. (B) Heatmap with enrichment values of different cancer types of the encounter classes on the trajectories. The encounter classes are ordered by eGFR values with kidney functions from healthy to impaired. (C) Correlation of enrichment values versus eGFR values for each cancer type across all trajectory classes. X-axis indicates the correlation, and the y-axis is the adjusted P-value. (D) Progression trajectories with enriched cancers. Color represents the corresponding TF-IDF score.

To further characterize the associations of different cancer enrichment with eGFR levels, we calculate their respective correlation with adjusted P-values shown in Figure 3C. Consistent with Figure 3B, GU cancer (cor = -0.884, adj. *P* = 9.25e-10) and skin cancer (cor = -0.885, adj. *P* = 9.04e-10) are shown to be highly and negatively associated with eGFR, with lower correlations than PRAC cancer (cor = -0.748, adj. *P* = 1.79e-5). Neuro cancer is positively correlated with eGFR (cor = 0.689), with a significant but higher adjusted *P*-value (adj. *P* = 0.0002). For these specific cancer types, their enrichment levels on the trajectory tree are visualized (Figure 3D). Indeed, skin cancer is shown with high enrichment values in classes with lower eGFR values and low enrichment levels in classes with higher eGFR values. Similar patterns are also observed in GU and PRAC cancer. An inverse pattern is observed for Neuro cancer, which is more enriched in high eGFR classes and less in low eGFR classes. The enrichment of other cancer types versus the kidney function is shown in Supplementary Figure 4.

### Certain cancers enriched in high-risk trajectories

With the associations between eGFR and age in different classes of the trajectory tree (Supplementary Figure 2C-D, Figure 4A), we identify two separately different trajectories, i.e., “high-risk” trajectory colored yellow and “low-risk” trajectory colored blue, that have lower correlations than the others. As mentioned above, the high-risk trajectory composed of classes 4, 19, 16, and 27 has lower eGFR in young people, indicating impaired kidney function. The low-risk trajectory, composed of classes 22, 28, and 26, has higher eGFR in elder people showing their healthy kidney function. Both high-risk, low-risk, and other trajectories show strong correlations between eGFR and age, demonstrating the reasonability of our learned progression trajectories. Figure 4B presents the different association patterns between eGFR and age in these two high-risk and low-risk trajectories specifically.

**Figure 4.**
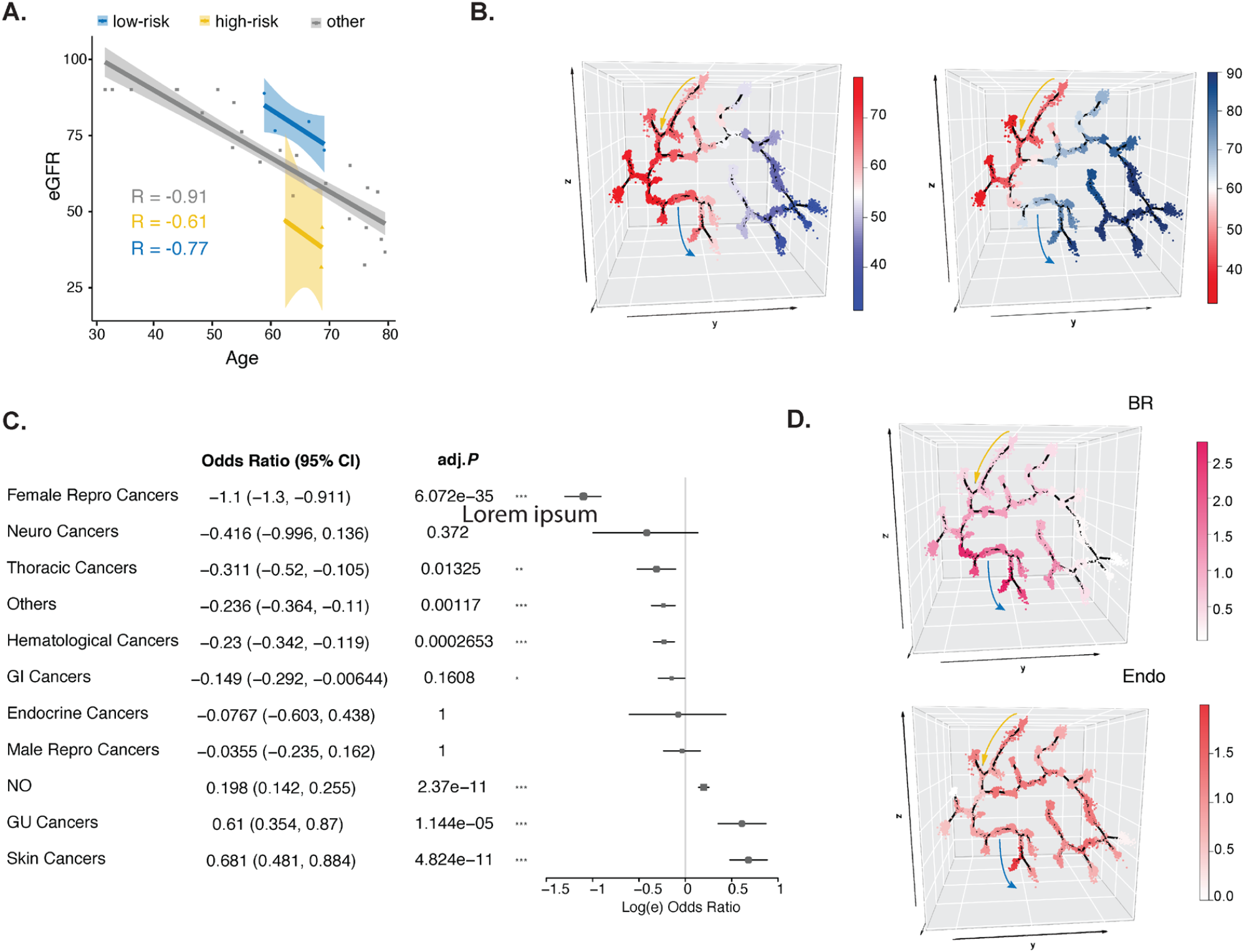
Identification of risk trajectories with different cancer enrichment. (A) The associations of eGFR and age in high-risk and low-risk trajectories. (B) Visualization of high-risk and low-risk trajectories of age and eGFR, denoted by yellow and blue arrows accordingly. (C) The forest plot shows the odds ratio and adjusted P-values of different cancer types between high-risk and low-risk trajectories. (D) Visualization of BR and Endo’s cancers enriched in high-risk and low-risk trajectories.

With these observations, we then investigate the odds ratios of different cancer types between high-risk versus low-risk trajectories. As shown in the forest plot (Figure 4C), we identify that Female Repro cancer (BR) has the largest odds ratio with a significant adjusted P-value between high-risk versus low-risk trajectory, representing that Female Repro cancer has more hazards in high-risk trajectory. Different from Female Repro Caner, GU and skin cancers have positive odds ratios indicating that they are more dominated in the low-risk trajectory. The visualization of this cancer enrichment at the high- and low-risk trajectories further verifies their consistent patterns (Figure 4D). Though the cancer information is not used in trajectory learning, patients with different cancers are still enriched in different trajectories. This cancer enrichment analysis strongly supports the validity of our learned trajectory, suggesting that trajectories learned from the EHR ECI data are capable of capturing kidney disease progression from different causes.

### Prediction of chronic disease progression

With the learned trajectory, our DEPORT approach can also effectively learn and predict new patient encounters in the progression trajectory. Specifically, each of the new patient’s encounters will first be projected into the latent space by the trained graph-AI embedding model, then mapped onto the learned trajectories using the centroid of *k*-nearest neighbors in the latent space. The prediction performance is shown in Figure 5A. Based on the trajectories of the learning cohort, we observed strong associations between eGFR and age (cor = -0.89) and less correlation for the high-risk (cor = -0.87) and low-risk (cor = -0.73) trajectories. Such associations are also captured in the prediction cohort (cor = -0.91), as well as in the high-risk (cor = -0.61) and low-risk (cor = -0.77) trajectories, respectively. These results demonstrate the superior prediction performance of our approach and the accurate progression pattern. Further investigation of the enriched Female Repro cancers (BR) in learning and prediction trajectories also suggests consistent results. Since patient numbers in EHR data can easily reach multiple million levels, it is promising that our DEPORT approach learns the consistent trajectory patterns in both learning and prediction cohorts, demonstrating its effectiveness and capability for large amounts of EHR data.

**Figure 5:**
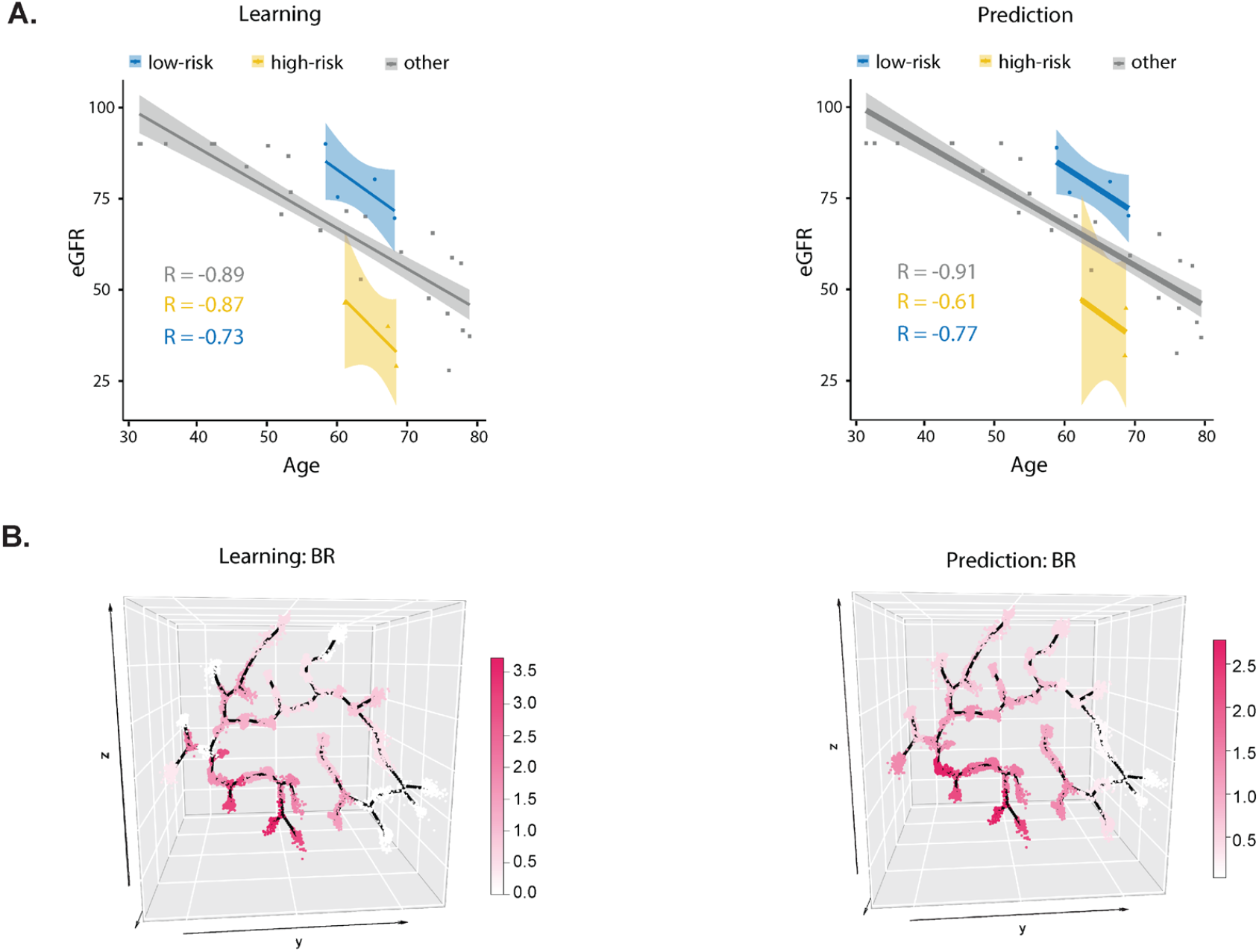
Prediction of new encounters on the progression trajectories. (A) The consistent associations of age and eGFR on the trajectories learned from the learning cohort and the prediction cohort. (B) Visualization of BR enrichment score in learning cohort and prediction cohort, respectively. Color represents the corresponding TF-IDF score.

## Discussion

The fragmentation and heterogeneity of EHR data are challenges to current longitudinal analytical and modeling approaches. On the one hand, missing values dominate EHR data. They are beyond meaningful data imputation, as the existence of a data value is the result of a real-world decision after negotiations between physicians, payers, and patients according to the best knowledge at that time, clinical need, socioeconomic status, policies of insurers and payers, cultural influence, and patient’s preference of healthcare providers. Moreover, clinical features are of various temporal patterns, which reflect the difference in reasons for clinical visits, health conditions, treatment outcomes, and follow-up plans. Many time series analytical tools, such as group-based trajectory modeling [16-19] and stochastic differential equations [20], are not directly applicable to such longitudinal data. In addition, current available CKD risk prediction models are of limited utility in patients without CKD [21]. These models require covariates such as eGFR that are validated for individuals with CKD. Identifying at-risk individuals in vulnerable populations and delivering them to providers early in a patient’s disease course is critical. Patients and providers can then make informed decisions on lifestyle modifications or alter deleterious medication regimens before irreversible damage is done to the kidney.

The graph-AI-based latent representation of heterogeneous and informatively censored clinical and molecular features is novel. It is attracting growing interest in biomedical research, including integrating real-world evidence data for precision oncology [22] and complex omics data for knowledge transfer across disparate datasets [23, 24]. This work uses a graph-based AI model to represent highly heterogeneous and fragmented longitudinal EHR data. With the learned representation, we then use the reversed graph embedding to learn disease progression trajectories from EHR data. As reversed graph embedding approach [6, 25, 26] assumes that the longitudinal data points are fragmental and sampled from the underlying trajectories (mathematically known as the principal graph underlying the observed fragmental data points), thus, it is intrinsically suitable for learning trajectories from temporally fragmental clinical observations. By extending our previous work [26] to systematically outline the progression landscape of CKD, our DEPORT approach casts new light on EHR-based clinical research. The DEPORT approach addresses the two major limitations of EHR data analysis and fully unleashes the power of the highly incomplete and informatively censored EHR data with heterogeneous features.

Taking together, our work suggests that: 1) the graph-AI-based latent representation captures the essential clinical and biological underpinnings of the heterogeneous CKD progression trajectories; 2) the age network captures the longitudinal patterns and the time information in EHR data for graph AI models; 3) the inductive representation learning model is robust and highly scalable, allowing efficiently learning trajectories from million-level data through random sampling. The capability of learning longitudinal associations between trajectories and potential causes in very large data is crucial in EHR data mining; 4) graph AI models can effectively address EHR challenges, including heterogeneous inputs, informatively censored features, non-linear effects, and low count inflation of less common events and clinical conditions. The graph-AI-based latent representation and reversed graph embedding are suitable to robustly learn CKD progression trajectories from different causes when the current CKD monitoring indexes failed.

## Data Availability

The DEPOT method is provided as an open-source package on GitHub (https://github.com/jing-su/depot).
The EMR data of the patients cannot be made available to the public.

## Acknowledgments

J.S. is supported by the National Library of Medicine of the National Institutes of Health (award number R01LM013771), the National Institute on Aging of the National Institutes of Health (award number R01LM013771-02S1), the National Cancer Institute (Indiana University Melvin and Bren Simon Comprehensive Cancer Center Support Grant P30CA082709), and Indiana University Precision Health Initiative. Q.S. is supported by the National Institute of General Medical Sciences of the National Institutes of Health (award number R35GM151089).

## Code availability

The DEPOT method is provided as an open-source package on GitHub (https://github.com/jing-su/depot).

## The role of the Institutional Review Board

The study has been covered under the Wake Forest School of Medicine Institutional Review Board (RB) and the Indiana University IRB-approved protocol with a waiver of the consent of the secondary use of deidentified EMR in research.

## Figure and table legends

**Table 1. Cohort demographic characteristics.**

**Supplement Figure 1.**
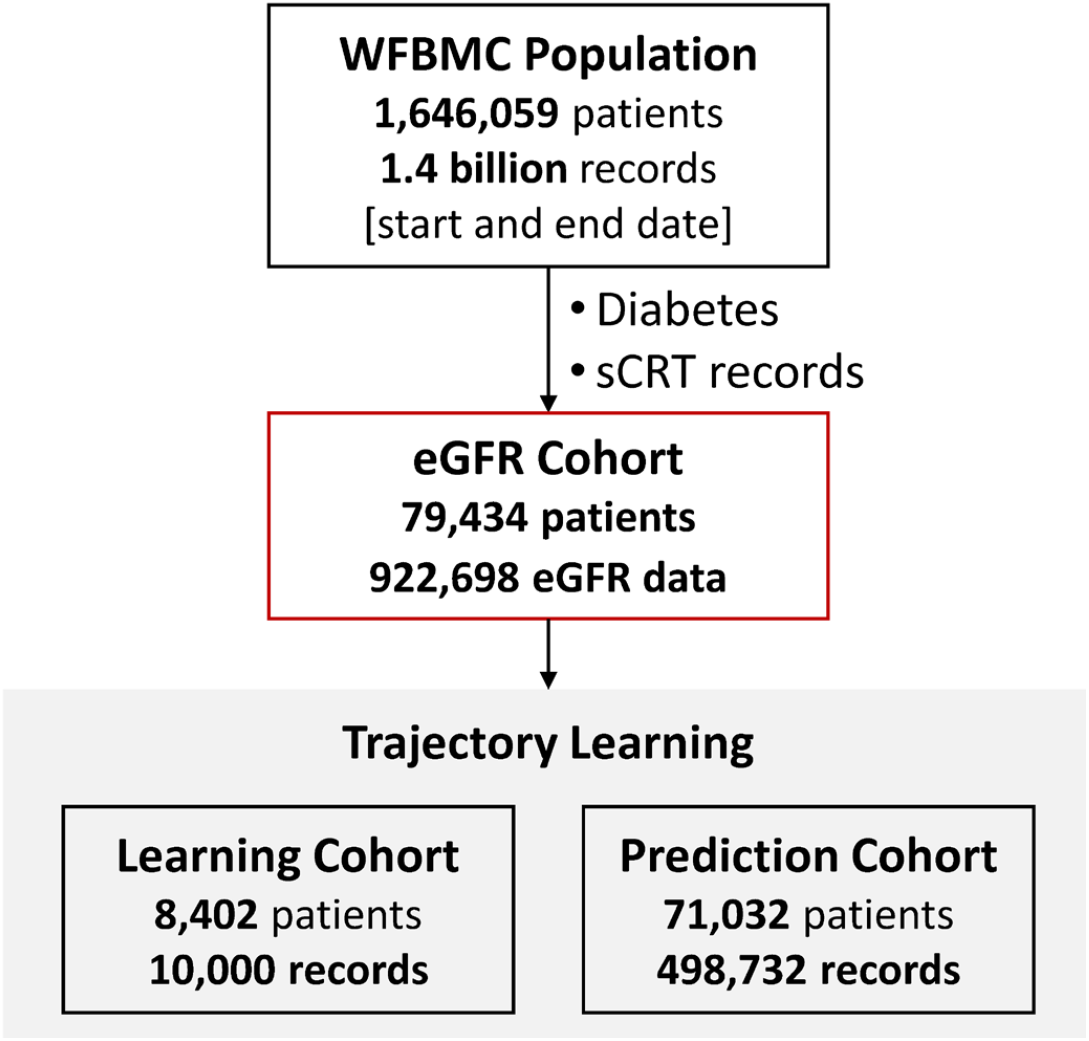
CONSORT diagram.

**Supplement Figure 2.**
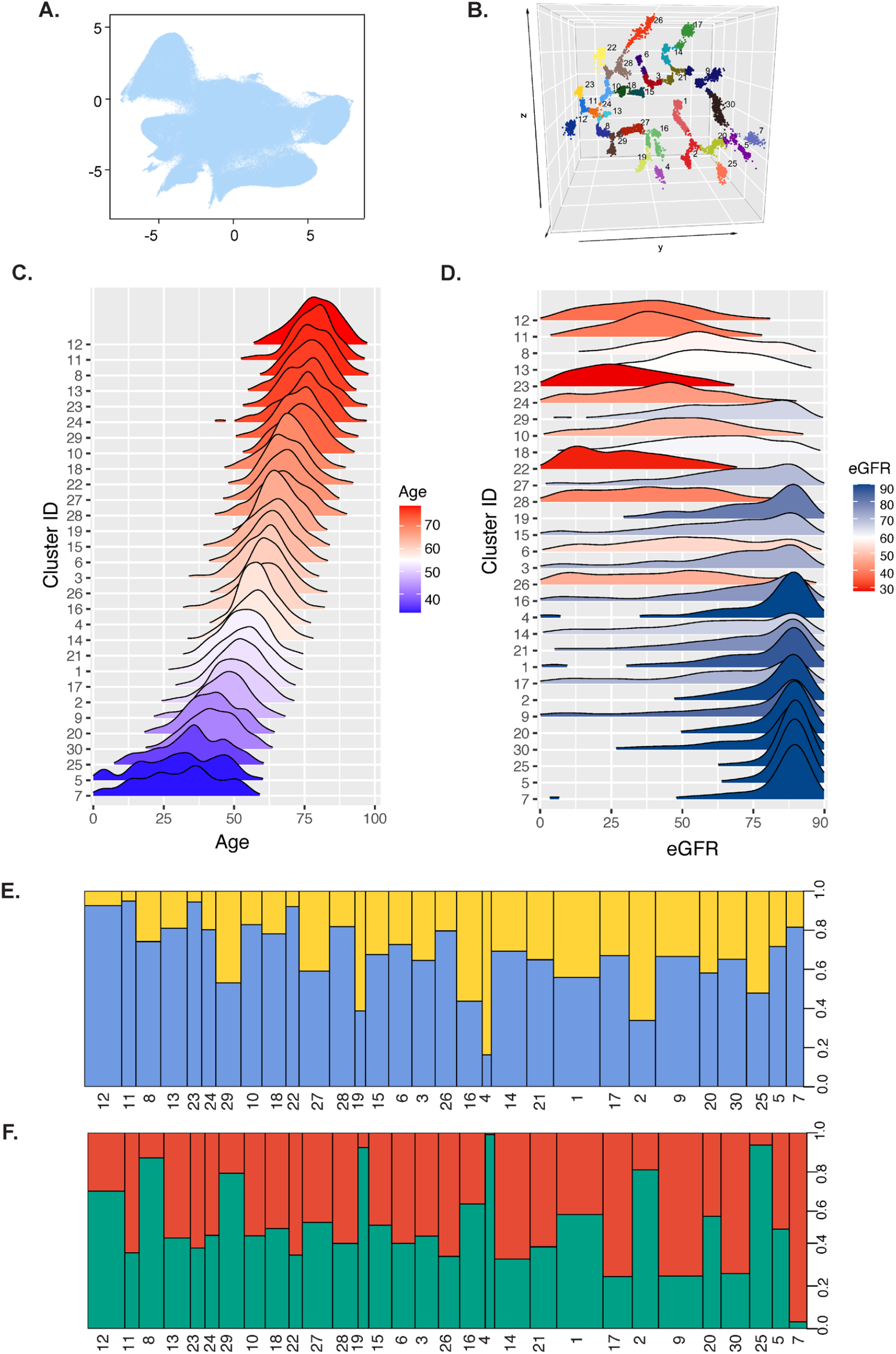
Representation space and the interpretation of class-based trajectories. (A) UMAP visualization of the representation space learned by the graph-AI model in our DEPORT approach. The representation space consists of encounters shown as dots. (B) Progression trajectories learned from the representation space. Encounters are shown as dots color-coded and labeled by different classes. (C) Trajectory classes are ordered by age. (D) eGFR values on trajectory classes ordered by age. (E) The proportion of race distributions on trajectory classes is ordered by age. (F) Proportion of sex distributions on trajectory classes ordered by age.

**Supplement Figure 3.**
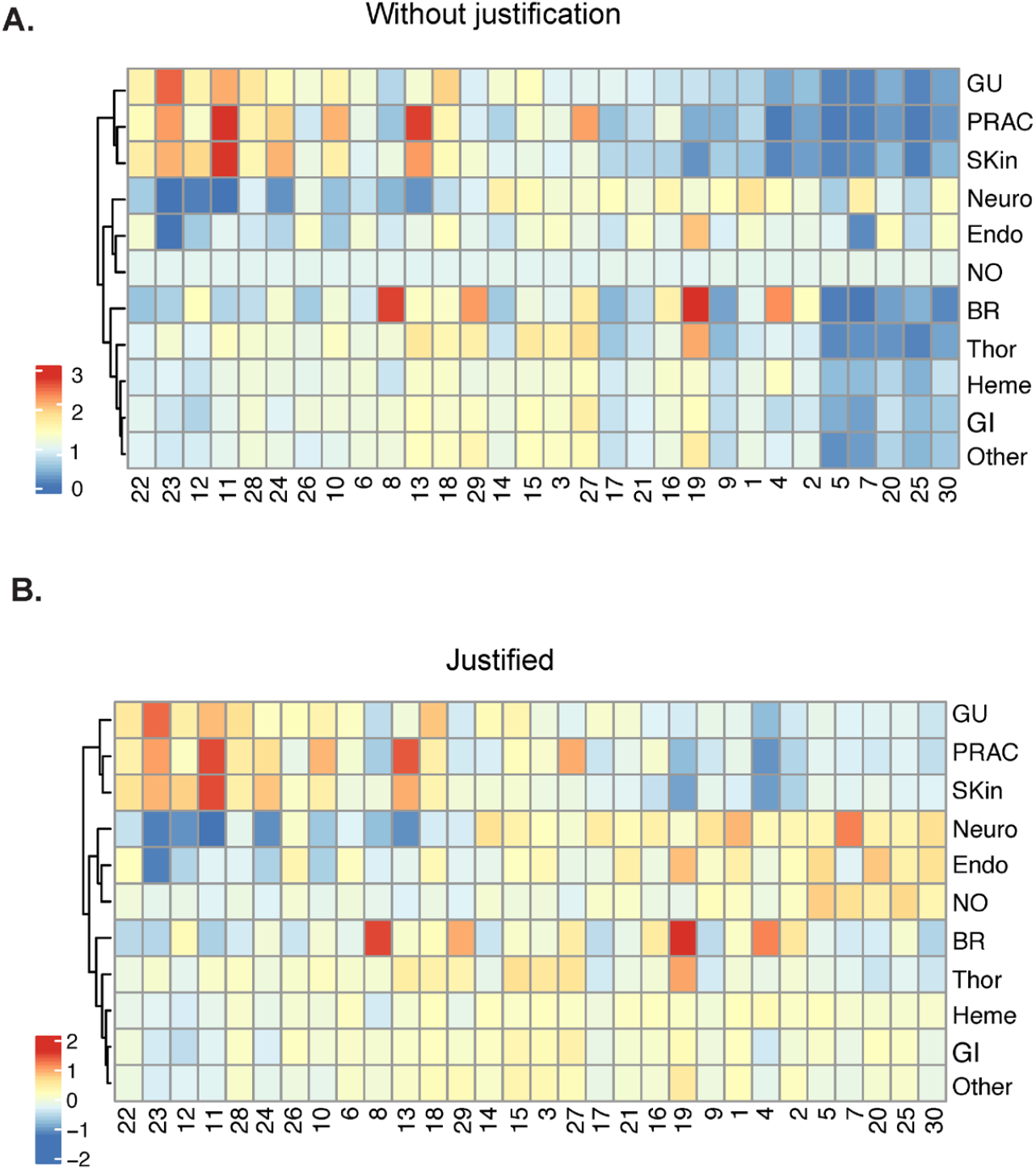
Cancer enrichment analysis on class-based trajectories. (A) Heatmap with enrichment values of different cancer types of the encounter classes on the trajectories. The encounter classes are ordered by eGFR values with kidney functions from healthy to impaired. (B) Heatmap with enrichment values of different cancer types after removing the confounding effects of age.

**Supplement Figure 4.**
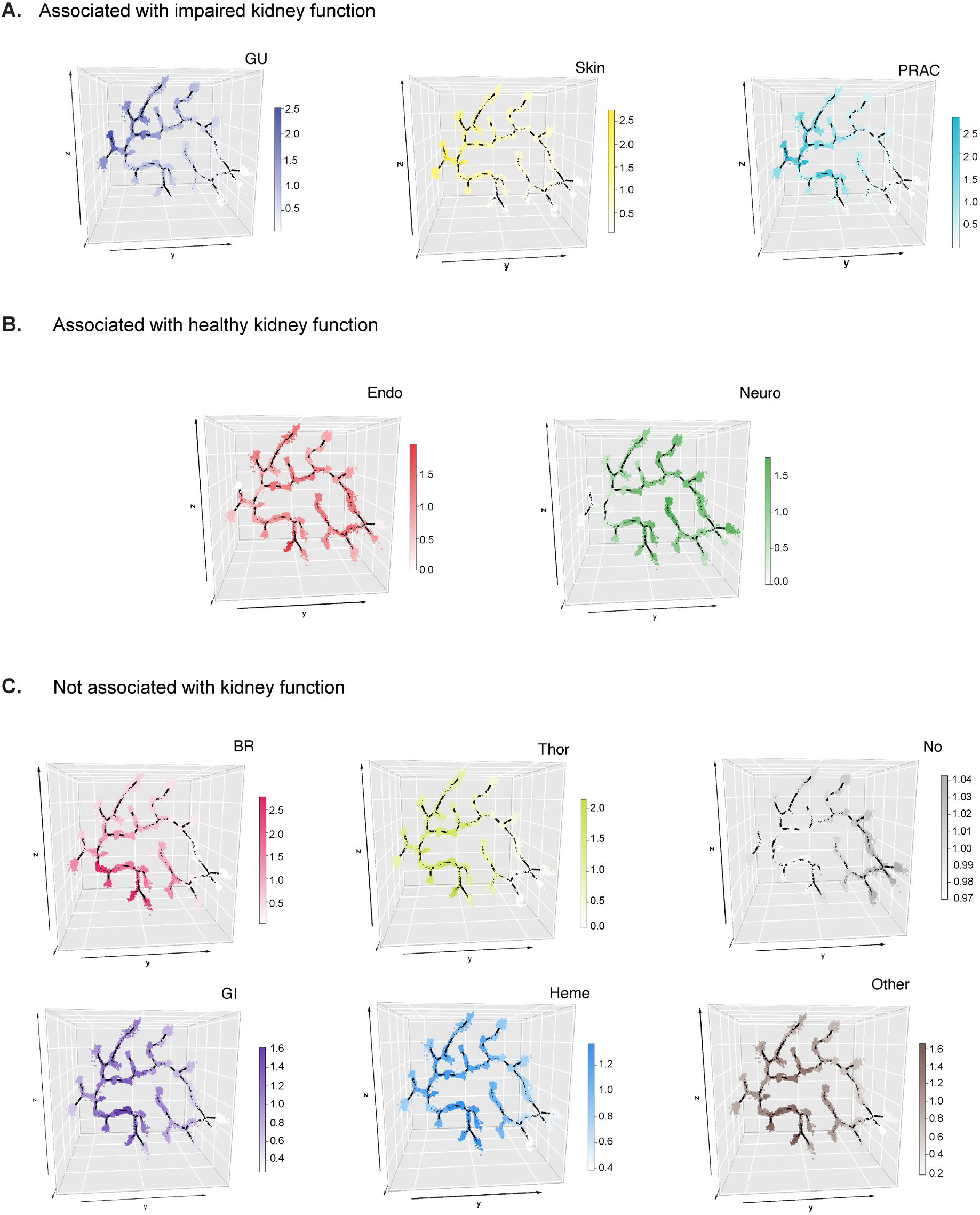
Enrichment of different cancer types on the trajectories. (A) Visualization of trajectories with enriched cancers that are associated with impaired kidney functions. (B) Trajectories with enriched cancers associated with healthy kidney functions. (C) Trajectories with enriched cancers that are not associated with kidney functions.

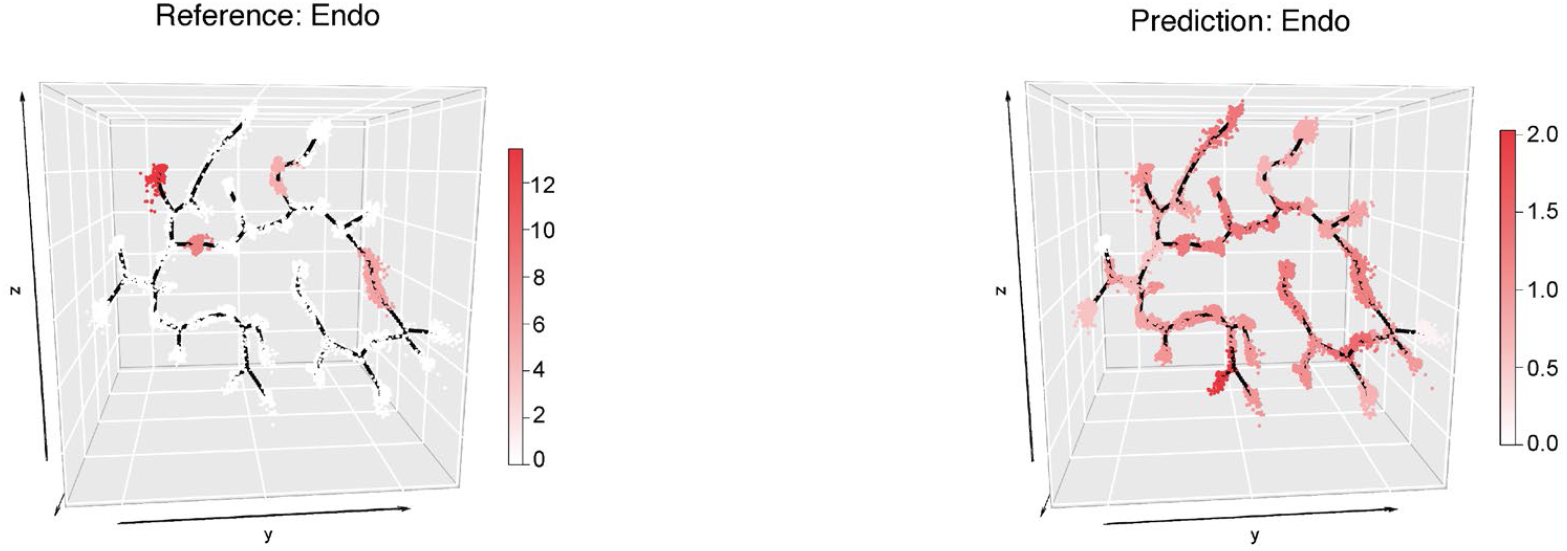

